# Designing national programs for expanded carrier screening: Results from a discrete-choice experiment in Singapore

**DOI:** 10.64898/2026.04.09.26350563

**Authors:** Robin Blythe, Sameera Senanayake, Yasmin Bylstra, Jack Roberts, Christina Choi, Myrabeth Juann Yeo, Jeanette Goh, Nicholas Graves, Ai Ling Koh, Saumya Shekhar Jamuar

## Abstract

**Background:** Carrier screening for inherited genetic disorders can reduce the burden of conditions that lead to childhood morbidity and mortality, including thalassaemia, cystic fibrosis, and spinal muscular atrophy. To be successful, national carrier screening programs should aim to maximise uptake, which may depend on population preferences for screening characteristics. In this study, we aimed to determine how expanded carrier screening in Singapore should be designed based on operational factors including suggested copayments, wait times, and disorders included in screening panels.

**Methods:** We elicited stated preferences for the design of a hypothetical national carrier screening program with seven attributes from 500 Singaporeans of reproductive age (18 to 54). A discrete choice experiment was applied using 30 choice tasks with 3 alternatives per task, divided between 3 blocks. The mixed multinomial logit model was used to estimate willingness-to-pay for each attribute level. Predicted uptake for three plausible screening programs was assessed, with copayment amounts from $0 to $1,200 in increments of $30. Impact on the annual national budget was calculated as a function of 25,000 expected eligible couples per year. All costs were reported in 2026 SGD.

**Results:** Respondents showed the strongest preferences for cost, followed by the number of diseases included in the panel, then wait times, with limited impact of remaining attributes. With no copayments, predicted uptake ranged from 85% [95% CI: 83% to 87%] to 90% [88% to 92%] for the basic and utility-maximising screening programs, respectively. This declined to 61% [56% to 66%] and 69% [65% to 73%] and, respectively, at a copayment of $1,200 per test. The model predicted higher uptake if a selection of screening alternatives were available, compared to a single program. The budget impact was highly dependent on population eligibility, copayments, and couples’ decision-making processes, but was unlikely to exceed $22.5m [$19.0m to $26.6m] per year unless expanded beyond married couples.

**Conclusions:** There was high predicted demand for carrier screening even as copayments increased. Successful strategies to improve uptake may include reducing copays and wait times, increasing the number of screening options available to prospective parents, and increasing program eligibility beyond pre-conception married couples.

## BACKGROUND

Autosomal recessive genetic disorders affect around 2% of births.^1^ While some heritable conditions only become apparent later in life, many, such as spinal muscular atrophy, lead to neonatal or childhood mortality, lifelong disability, and cognitive impairment in the absence of treatment.^2^

Carrier screening programs for couples prior to conception can reduce the rate of births affected by genetic disorders including cystic fibrosis^3^ and thalassaemia.^4,5^ During carrier screening, both prospective parents provide DNA samples obtained from blood, saliva, or buccal swabs. Genetic sequencing identifies disease-causing variants in genes associated with inherited conditions. For autosomal recessive conditions, if both parents are carriers of a relevant variant in the same gene, they are at increased risk as there is a 1 in 4 chance of having a child affected by that condition.

Rapid reductions in sequencing costs make it feasible to add a variety of disorders to screening panels^6^ to provide carrier screening at scale to the general population. A national carrier screening program for disorders with high rates of childhood morbidity and mortality has already been successfully implemented in Israel,^7^ and a pilot program is underway in Singapore to reduce the burden of genetic disorders.^8^

To tailor a national carrier screening program for Singapore, policymakers may weigh the program characteristics that encourage uptake and effectively reduce the number of affected births while minimising budget impact. Selecting conditions to include is not straightforward; genomics data have historically featured European populations, meaning that disorders with a higher prevalence in Singapore may be underrepresented.^9,10^ Operational considerations, including wait times, counselling availability, and copayment amounts can all affect program uptake, thereby limiting screening rates.

By eliciting the preferences of the general population for carrier screening, we aim to illustrate how expanded carrier screening can move from pilot to policy in Singapore. We report the results of a discrete-choice experiment among the general population of Singaporeans of reproductive age to determine how a national expanded screening program should be designed to maximise uptake without incurring high operational costs.

## METHODS

### Study overview

We conducted a systematic review and meta-analysis to identify potential attributes and levels for the choice task. Results were localised to Singapore based on expert advice, and volunteers performed a think-aloud exercise to validate whether respondents would understand the meaning of each attribute and level. We conducted a pilot study to confirm preference consistency among participants before releasing the full survey to achieve our target sample size. The study flow diagram is shown in Figure 1, and the final survey design is shown in the Supplement, Appendix 1. This study was approved by the SingHealth Centralised Institutional Review Board (CIRB2019/2243).

**Figure 1:**
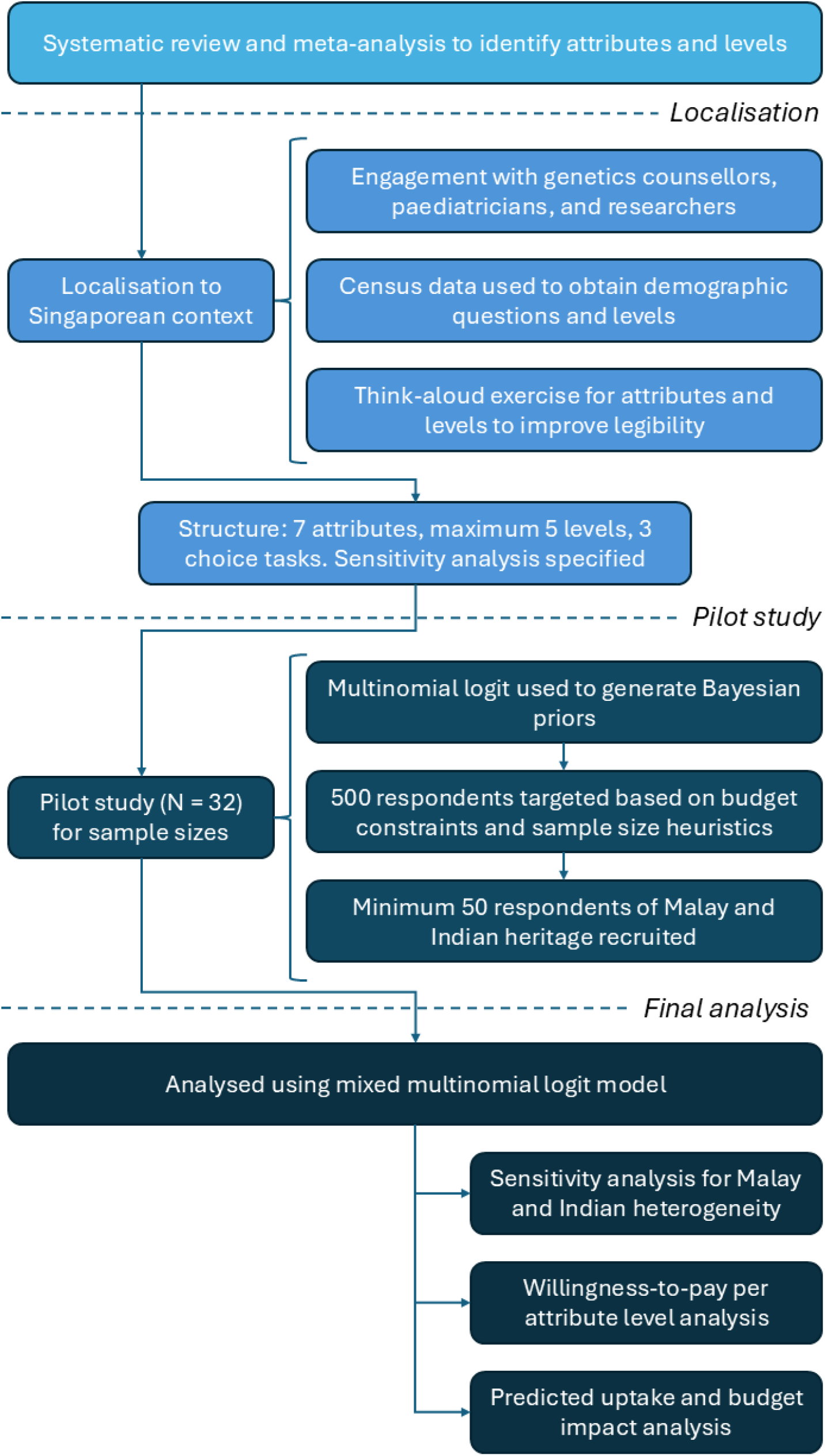
Flowchart of the study design and recruitment process

### Experiment design

Prior to this study, we conducted an integrative review and meta-analysis to determine attributes that should potentially be included in our experiment.^11^ Briefly, we identified seven distinct attributes: 1) the type of healthcare provider to oversee screening; 2) the format of screening education and counselling that would be received by the couple (e.g., in-person or online); 3) the screening method (e.g., stepwise or couple screening); 4) the types of conditions included in screening; 5) when screening should be available (e.g., pre-conception or at any time); 6) waiting times for results; and 7) willingness-to-pay. We refer interested readers to Yeo Juann et al (2026). Attribute and level localisation to the Singaporean context is described in the Supplement, Appendix 2.

We constructed the choice tasks using an experimental design. Each task presented respondents with two hypothetical screening program alternatives and an opt-out option. The two unlabelled alternatives were presented as ‘Screening program A’ and ‘Screening program B’. The inclusion of an opt-out alternative was intended to enhance the policy relevance of the choice tasks by allowing us to predict program uptake. The final choice task included seven attributes, which were incorporated into a fractional factorial design using Ngene software to generate a D-efficient design.^12^ The final experiment design comprised of 30 choice tasks, divided into three blocks of ten choice tasks per respondent.

A market research company, Kantar,^13^ was engaged to distribute the survey in English to Singaporeans aged between 18 and 54. A pre-screening question was included to exclude participants who believed carrier screening should not be available at all. A pilot survey (N = 32) was first distributed to validate the experiment design. Data from the pilot were analysed using a multinomial logit model, and the resulting parameter estimates were used as priors for the main experiment based on a Bayesian D-efficient design. Initial sample size calculations were based on statistical significance, and suggested a sample of over 15,000 respondents. As this was not feasible, we instead based our sample size on a combination of budget constraints and practical heuristics,^14^ with a final sample of 500 respondents. To ensure we obtained a sample roughly corresponding to the Singaporean population, we specified at least 50 respondents each identifying as Malay and Indian, who comprise 13.5% and 9.0% of the Singapore population aged between 20 and 54, respectively (Supplement, Appendix 3).

### Statistical and policy analysis

We used a mixed multinomial logit model to assess potentially heterogeneous preferences among study participants. The reference category was selected based on the most basic service provided: level 1 for how testing was conducted and the type of condition screened, and level 3 for the remaining attributes (Table 2, below). As we presented survey respondents with two screening alternatives with varying attribute levels plus an opt-out, (three alternatives per choice task), we specified alternative 3 (opt-out) as the alternative-specific constant (ASC). We specified cost as a continuous variable, and included all non-reference responses as both fixed and random effects, with random effects specified as normally-distributed.

Model convergence was assessed by iteratively increasing the number of Sobol draws from 200 in increments of 150, considering the model converged when the coefficients changed by less than 0.02 between model fits. To check whether a mixed model specification was supported by the data, we applied a likelihood ratio test and checked the Akaike Information Criterion (AIC) between models with and without random effects specifications.

Finally, we estimated willingness-to-pay for each attribute by dividing the attribute utility effect sizes by the effect size for cost. We predicted program uptake under three hypothetical policies reflecting the reference category (Policy 1), the program most similar to the ongoing pilot study (Policy 2) and a program that sought to maximise predicted utility for the lowest additional service costs (Policy 3). Briefly, Policy 2 was similar to the reference category except it included both extremely severe and severe diseases, had shorter wait times, and included at least one counselling session for positive tests. Policy 3 included extremely severe, severe, and moderate conditions, was available to all, was delivered in a stepwise fashion, had 4-week wait times, and made counselling available both before and after testing. These are further described in the Supplement, Appendix 4.

As micro-costing was not possible, we used probabilistic estimates for uncertain service costs to estimate the plausible ranges of costs per policy component under each combination of policies, and summarised these simulated costs per couple to estimate budget impacts. Monte Carlo sampling with 10,000 iterations was used to sample uncertain input values. Costs were reported in 2026 SGD.

The model and model predictions are described in further detail in the Supplement, Appendix 4, and R code is provided in the online repository. Ngene software was used to design the experiment, but all further analysis, interpretation and visualisation was conducted in R,^15^ using the *logitr* and *ggplot2* packages.^16,17^

### Sensitivity analysis

As Singapore primarily consists of three relatively distinct self-reported ethnic groups (Chinese, Malay, and Indian) with potentially differing attitudes to government-sponsored programs, we pre-specified a sensitivity analysis to examine sensitivity to cost and the opt-out decision among Malays and Indians, leaving Chinese respondents as the reference category. The sensitivity analysis included interaction terms for: cost and Malay race; cost and Indian race; ASC and Malay race; and ASC and Indian race. We assessed model fit compared to the main model using the likelihood ratio test and AIC.

### Data sharing and code availability

All model code and anonymised survey results are available at https://github.com/robinblythe/DCE_pop_screen.

## RESULTS

Respondents mostly self-reported as ethnically Chinese, with Buddhism or no affiliation to any religion being the most commonly reported. Race and religion broadly aligned with Singapore census results. Most respondents held a university degree; two-thirds of our sample held at least a bachelors, compared to just one-third of the general population, and we only received 1% of responses from respondents who did not finish secondary school, in contrast to 24% of the population. Our sample also contained a larger proportion of unmarried respondents compared to the general population, and 60% reported either planning to have children or being unsure. Finally, 71% of our sample reported knowing no one with an inherited condition confirmed by genetic testing. Respondent characteristics are shown in Table 1.

**Table 1:** Respondent characteristics relative to census data. Our sample inclusion criterion was aged 18 to 54, while census data reports age from 20 to 54.

| Demographic variable | Summary statistics | Singapore population statistics |
| --- | --- | --- |
| Age | Mean 37.2 (SD 10.1) |  |
| Gender* |  |  |
| Female | 249 (49.8%) | 1,795,534 (51.9%) |
| Male | 249 (49.8%) | 1,663,560 (48.1%) |
| Non-binary/third gender | 1 (0.2%) | Unknown |
| Prefer not to say | 1 (0.2%) | Unknown |
| Ancestry** |  |  |
| Chinese | 378 (75.6%) | 3,109,075 (74.0%) |
| Malay | 50 (10.0%) | 569,134 (13.5%) |
| Indian | 50 (10.0%) | 379,098 (9.0%) |
| Other | 22 (4.4%) | 147,208 (3.5%) |
| Religious affiliation* |  |  |
| Buddhist | 160 (32.0%) | 1,074,159 (31.1%) |
| No religious affiliation | 99 (19.8%) | 692,528 (20.0%) |
| Islam | 52 (10.4%) | 539,251 (15.6%) |
| Other Christian | 75 (15.0%) | 411,674 (11.9%) |
| Taoist | 35 (7.0%) | 303,960 (8.8%) |
| Catholic | 38 (7.6%) | 242,681 (7.0%) |
| Hindu | 31 (6.2%) | 172,963 (5.0%) |
| Other | 10 (2.0%) | 21,878 (0.6%) |
| Highest level of education completed* |  |  |
| PhD or Masters degree | 46 (9.2%) | 1,007,183 (32.1%)<sup>†</sup> |
| Bachelors degree | 275 (55.0%) |  |
| Non-university higher education (e.g., polytechnic) | 121 (24.2%) | 861,819 (27.4%) |
| Secondary school | 56 (10.6%) | 505,564 (16.1%) |
| Did not finish secondary school | 5 (1.0%) | 765,869 (24.4%) |
| Marital status* |  |  |
| Single | 183 (36.6%) | 775,711 (24.7%)<sup>†</sup> |
| In a relationship | 44 (8.8%) |  |
| Married | 254 (50.8%) | 2,031,836 (64.7%) |
| Divorced/widowed/separated | 19 (3.8%) | 332,889 (10.6%) |
| Currently pregnant | 60 (12.0%) | Unknown |
| Have children | 228 (45.6%) | Estimated at 46.5%<sup>‡</sup> |
| Planning on having children in future |  |  |
| Yes | 207 (41.4%) | Unknown |
| No | 198 (39.6%) | Unknown |
| Unsure | 95 (19.0%) | Unknown |
| Do you or someone you know have an inherited genetic condition confirmed by genetic testing (e.g., thalassaemia)? |  |  |
| Yes, I do | 59 (11.8%) | Unknown |
| Yes, my children do | 12 (2.4%) | Unknown |
| Yes, someone I know does | 73 (14.6%) | Unknown |
| No, not to my knowledge | 356 (71.2%) | Unknown |

\*Based on 2020 census results, last updated 16/06/2021
\*\*Singapore Department of Statistics data extract, last updated 29/09/2025
<sup>†</sup>The Singapore census does not differentiate between these groups
<sup>‡</sup>Estimated based on resident households by living arrangement in 2024, unadjusted for age.

### Multinomial logit results

The average marginal utility of each attribute relative to the reference category after accounting for unobserved heterogeneity is represented in Table 2. Respondents primarily expressed preferences for: being able to test at any time, including before marriage; including more conditions in the screening panel; having the opportunity for counselling both before and after testing; and shorter waiting times. Cost was a strong predictor of uptake. There was also a large negative coefficient for the ASC (- 5.1), indicating that respondents felt they would need to be compensated to forego screening entirely. Willingness-to-pay per attribute level is illustrated in Figure S1 (Appendix 4).

**Table 2:**
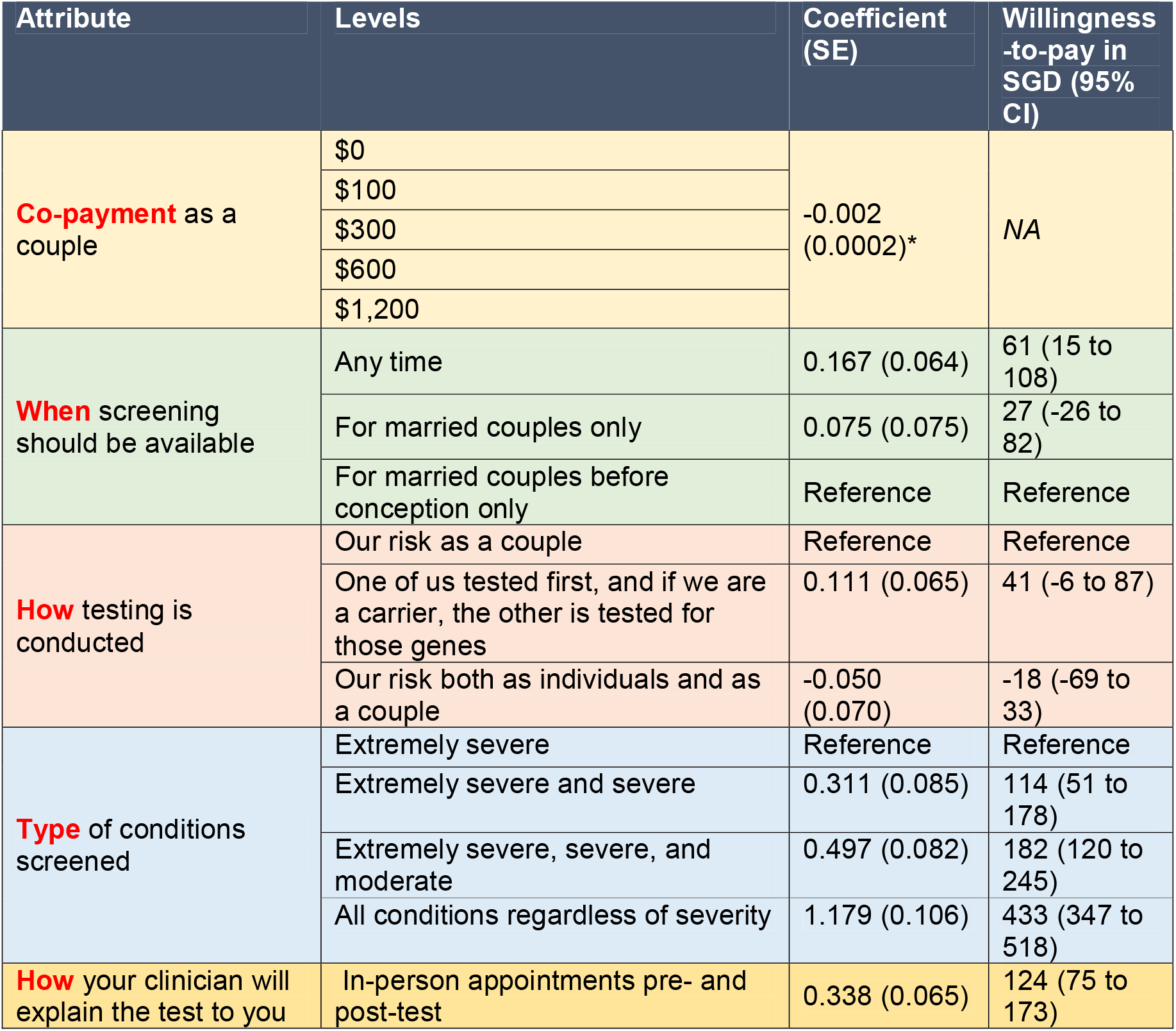
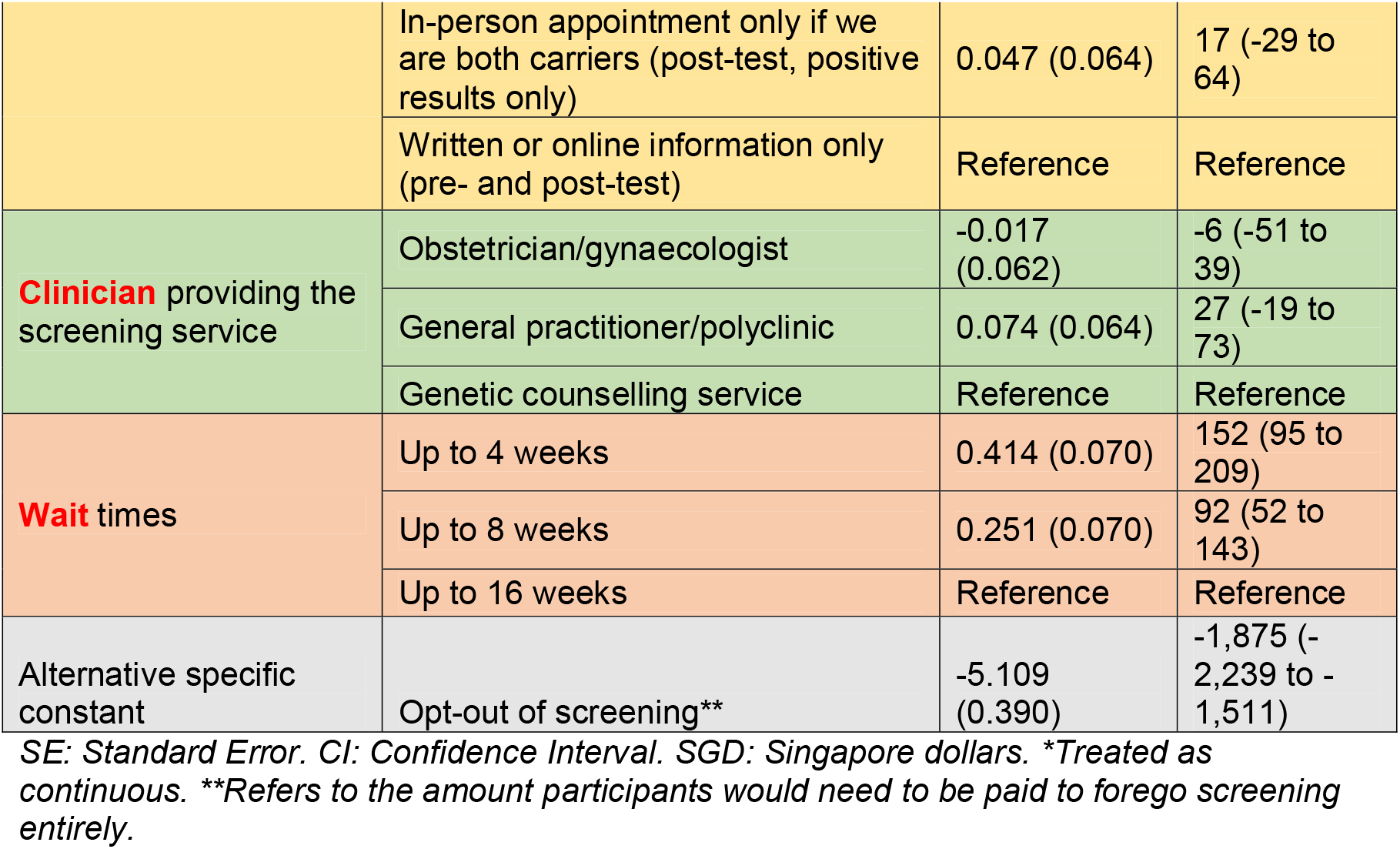
Results of the mixed multinomial logit model and willingness-to-pay for each attribute level after accounting for unobserved preference heterogeneity. Type of conditions screened was explained to participants as follows: Extremely severe (conditions with shortened lifespan in infancy/childhood or intellectual disability); Extremely severe and severe (conditions with shortened lifespan in early adulthood, impaired mobility, or disabling organ impairment); Extremely severe, severe, and moderate (conditions causing visual or hearing impairments and immune deficiency); All conditions regardless of severity, including conditions with onset later in life such as genetic risks of heart disease or cancer.

| Attribute | Levels | Coefficient (SE) | Willingness -to-pay in SGD (95% CI) |
| --- | --- | --- | --- |
| **Co-payment** as a couple | \$0 | -0.002 (0.0002)* | NA |
| | \$100 | | |
| | \$300 | | |
| | \$600 | | |
| | \$1,200 | | |
| **When** screening should be available | Any time | 0.167 (0.064) | 61 (15 to 108) |
|  | For married couples only | 0.075 (0.075) | 27 (-26 to 82) |
|  | For married couples before conception only | Reference | Reference |
| **How** testing is conducted | Our risk as a couple | Reference | Reference |
|  | One of us tested first, and if we are a carrier, the other is tested for those genes | 0.111 (0.065) | 41 (-6 to 87) |
|  | Our risk both as individuals and as a couple | -0.050 (0.070) | -18 (-69 to 33) |
| **Type** of conditions screened | Extremely severe | Reference | Reference |
|  | Extremely severe and severe | 0.311 (0.085) | 114 (51 to 178) |
|  | Extremely severe, severe, and moderate | 0.497 (0.082) | 182 (120 to 245) |
|  | All conditions regardless of severity | 1.179 (0.106) | 433 (347 to 518) |
| **How** your clinician will explain the test to you | In-person appointments pre- and post-test | 0.338 (0.065) | 124 (75 to 173) |
|  | In-person appointment only if we are both carriers (post-test, positive results only) | 0.047 (0.064) | 17 (-29 to 64) |
|  | Written or online information only (pre- and post-test) | Reference | Reference |
| **Clinician** providing the screening service | Obstetrician/gynaecologist | -0.017 (0.062) | -6 (-51 to 39) |
|  | General practitioner/polyclinic | 0.074 (0.064) | 27 (-19 to 73) |
|  | Genetic counselling service | Reference | Reference |
| **Wait times** | Up to 4 weeks | 0.414 (0.070) | 152 (95 to 209) |
|  | Up to 8 weeks | 0.251 (0.070) | 92 (52 to 143) |
|  | Up to 16 weeks | Reference | Reference |
| Alternative specific constant | Opt-out of screening** | -5.109 (0.390) | -1,875 (-2,239 to -1,511) |

SE: Standard Error. CI: Confidence Interval. SGD: Singapore dollars. \*Treated as continuous. \*\*Refers to the amount participants would need to be paid to forego screening entirely.

There was substantial preference heterogeneity for cost, test eligibility (whether unmarried or preconception couples were eligible), the most comprehensive list of disorders available, and 16-week wait times. While the average utility of testing, measured by the effect size of the ASC, was high (−5.11), the standard deviation of the random effect of the ASC was also high (−4.57), suggesting strong but heterogenous preferences for screening availability. After accounting for unmeasured heterogeneity, the model with interaction terms was not justified based on the likelihood ratio test and AIC. Estimates of the full model and sensitivity analysis, including heterogeneity, are shown in the Supplement (Appendix 4).

### Predicted uptake and impact on national budget

When presented with a choice between screening and opting out, the model predicted that uptake was highest for the utility-maximising program, and lowest for the basic screening program. With no copayment, predicted uptake was 90% [95% CI: 88% to 92%] and 85% [83% to 87%], respectively. These dropped to 69% [65% to 73%] and 61% [56% to 66%], respectively, at a copayment of $1200 per test.

However, when presented with a variety of options, overall uptake was predicted to increase to 93% [90% to 94%] when copayment was $0, dropping to 73% [69% to 76%] when copayment was $1,200. These results are shown in Figure 2. Conditional increases in program uptake, holding other predictors constant, is shown in Figure 3.

**Figure 2:**
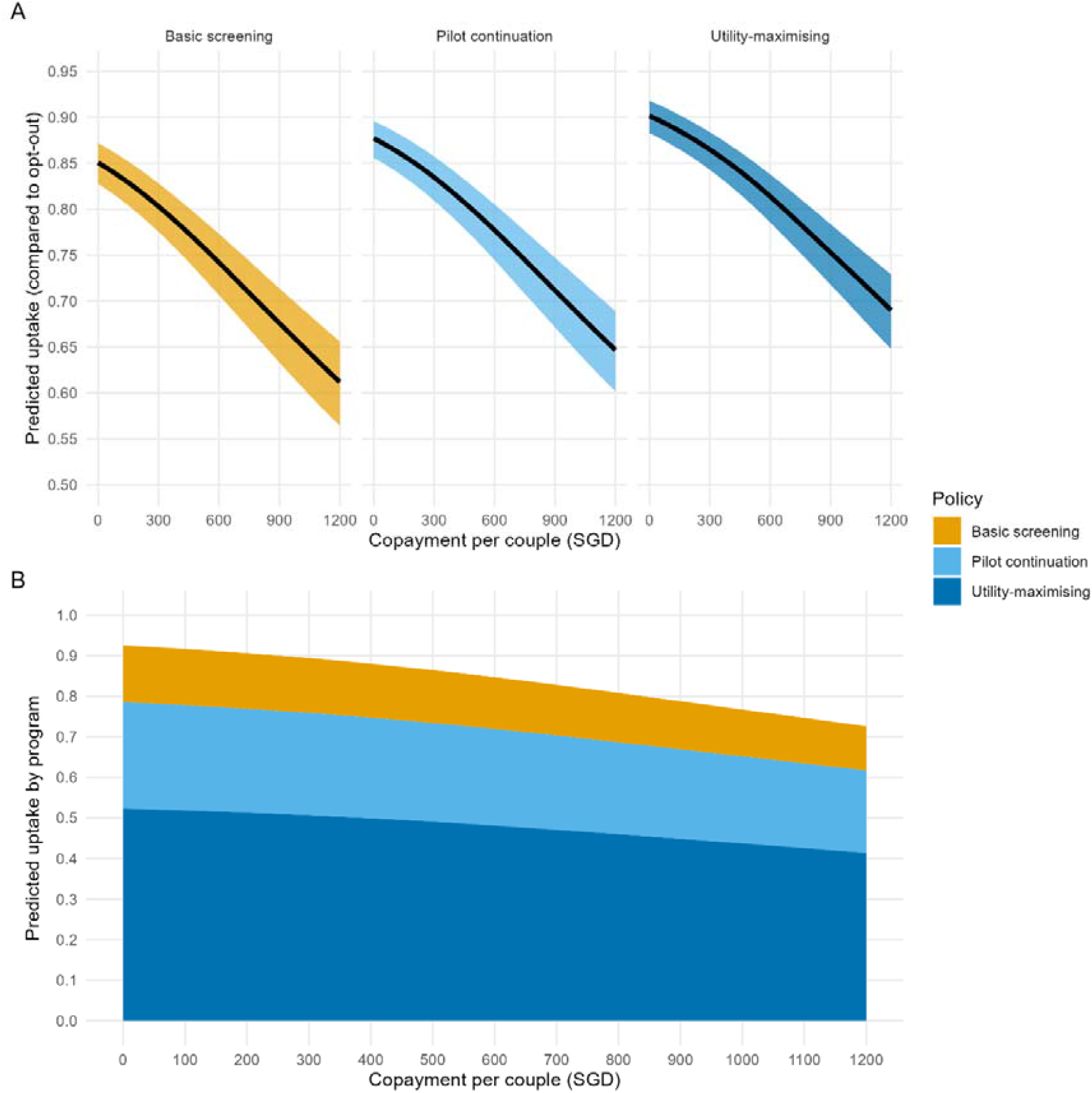
Estimated uptake as a function of test copayments when individual policies were compared to the opt-out (panel A) or offered as a menu of potential screening choices (panel B). Estimates assumed that an individual’s response was reflective of couple’s decision-making.

**Figure 3:**
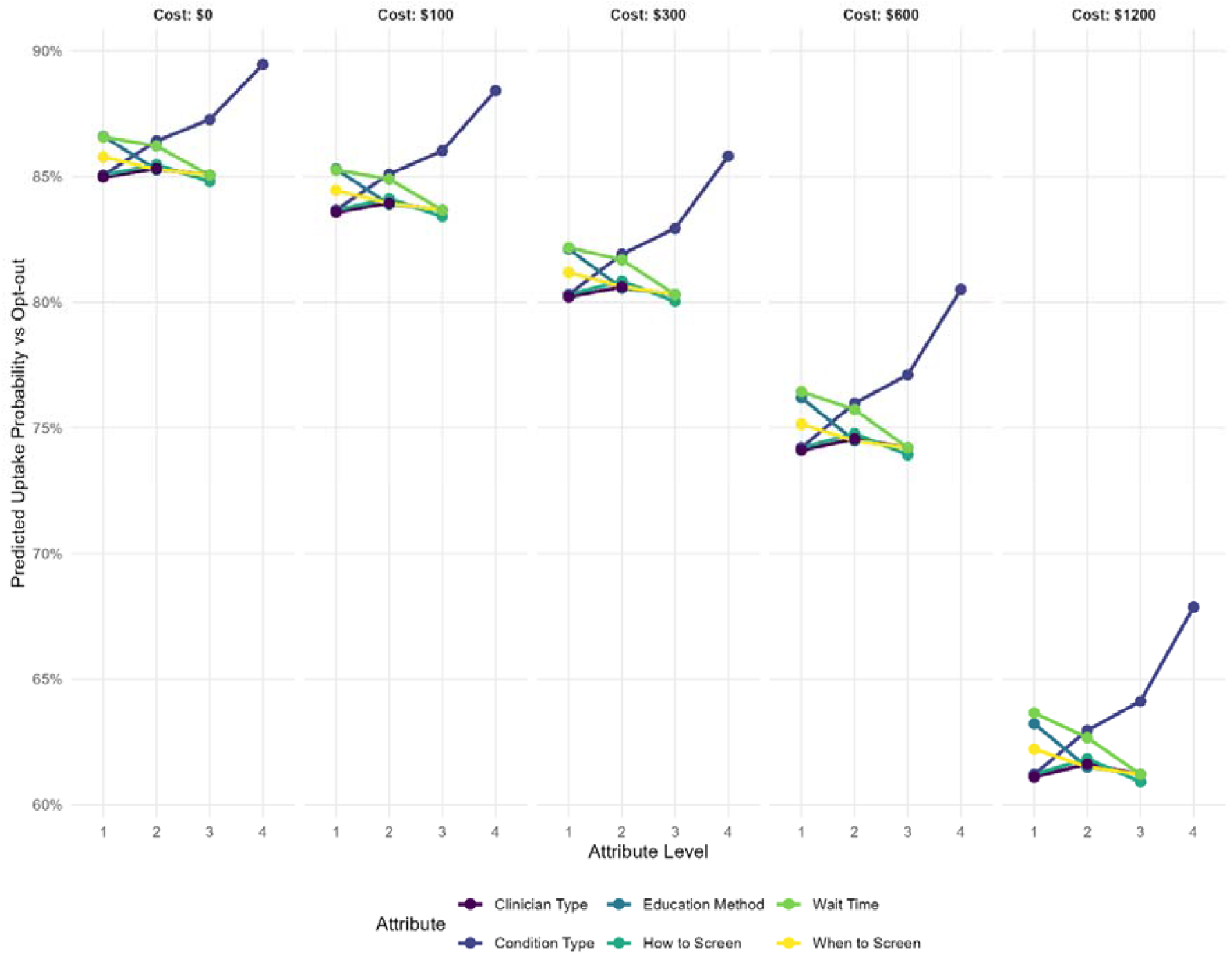
Predicted uptake versus opting out at each copayment amount as a function of the marginal value of each coefficient, with all other coefficients set to the reference category.

Budget impact was dependent on service costs, copayment amounts, and the decision-making model (individual or shared). Under a system with no copayment, assuming individual decision-making, budget impact was predicted to $7.8m [$5.7m to $10.1m] for the basic program, $18.9m [$18.4m to $19.4m] for extending the pilot program, and $22.5m [$19.0m to $26.6m] for the utility-maximising program. Budget impact reached $0 as copayments reached $450, $870, and $1,020 per couple for the basic, pilot, and utility-maximising programs, respectively. Assuming a menu of alternatives were available, the total cost was estimated at $20.2m [$15.9m to $25.4m] with no copayments. Figure 4 illustrates program uptake under single-program (panel A) and multiple program (panel B) policies.

**Figure 4:**
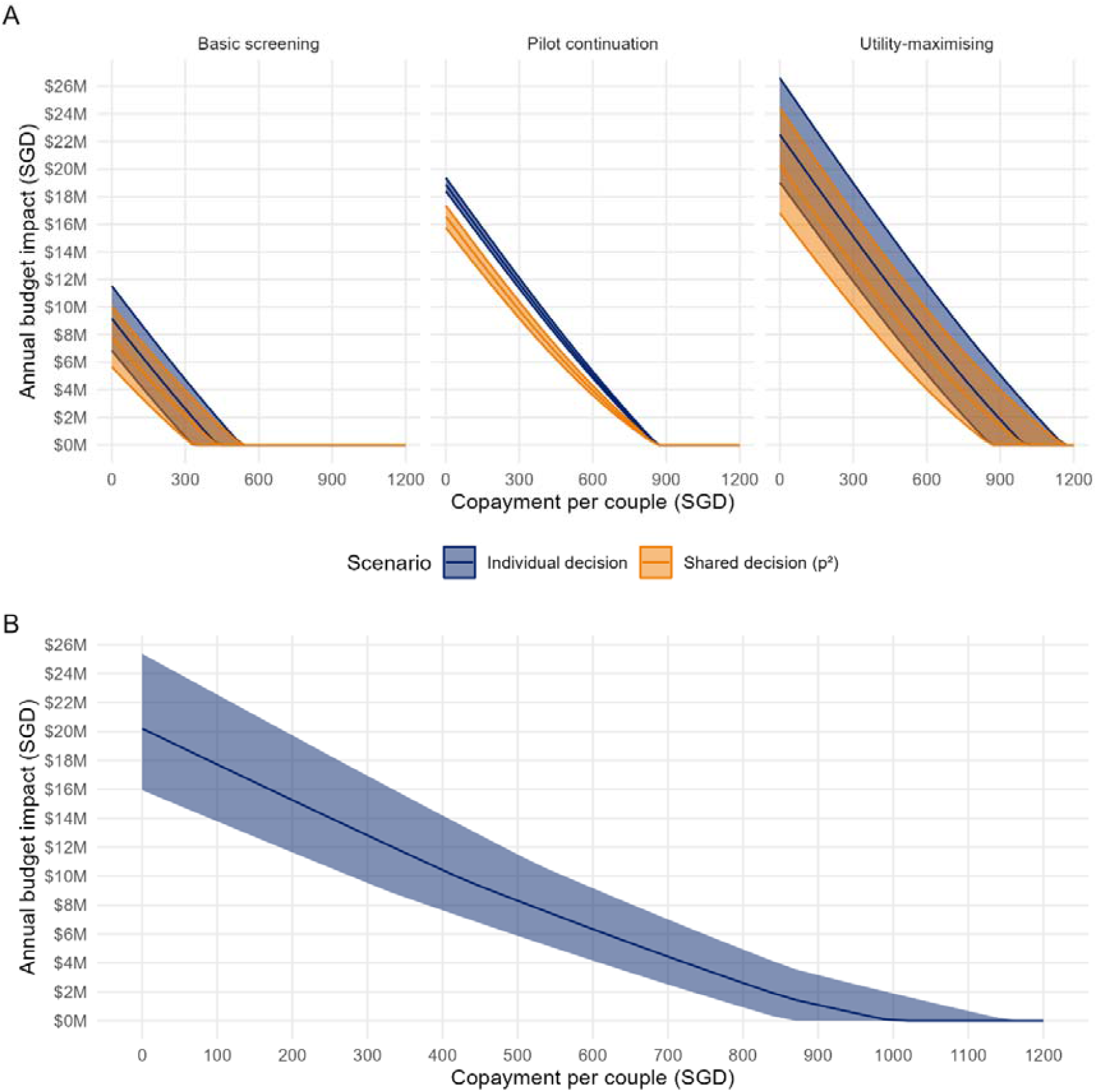
Estimated budget impact under individual and shared decision-making models for each policy (panel A) compared to a menu of potential screening options (panel B). Panel B assumes the individual decision-making model, as it was not feasible to model shared decision-making when multiple options were available. Budget impact assumed 25,000 couples per year would choose to undergo testing.

## DISCUSSION

Our findings suggest that there is strong demand for a localised carrier screening program in Singapore, and the model predicted that a large proportion of the population is likely to opt in to screening if it is perceived as being accessible and affordable, with short wait times. Respondents were most sensitive to price, the number of disorders screened, and wait times, but generally wanted the option to screen, indicated by the large negative coefficient on the ASC. Respondents may prefer screening programs conducted in a primary care setting and that consist of stepwise screening modalities, but these coefficient estimates had large error terms. Preferences similar to these have been previously observed in research of other screening program contexts,^11^ and were found to be relatively minor contributors to the overall utility of hypothetical screening programs.

We estimate that for a free screening program, after accounting for an estimated 5% of the population who believes screening should not be available at all, our model predicted that around 85% of the population could choose to participate. To maximise uptake, a selection of screening programs could be available, which would also reduce expected costs compared to only providing the utility-maximising program. Even under the most expensive program, annual costs were unlikely to exceed $26m based on our simulated conditions, and these costs could be reduced by the copayment mechanism, which both offsets health system expenditures and persuades couples to opt out.

### Policy relevance and recommendations for decision-making

Discrete choice experiments elicit stated preferences, which may predict, but not conform exactly, to decision-making. Prior research has shown that participants might overstate their intended participation;^18^ we posit that in the case of carrier screening, this may be due to several important differences between our experiment and clinical practice. First, there is a significant difference in effort required between stating one may be interested in screening on an online survey and actually following up. Reducing friction in the process of taking up screening by ensuring clinicians provide patients with adequate support may be an important facilitator.^19^ Second, our population was more likely to be unmarried and older than a typical adult seeking pre-conception screening. Preliminary results from our pilot program suggest that many couples may only seek out screening once they are beginning to attempt conception, after which carrier screening may be too late to affect decision-making. This will likely affect tolerance for wait times, especially as couples become pregnant. Accordingly, we recommend that screening be promoted early enough that it can provide meaningful information for couples planning for a family.^11^

The structure of a national screening program will depend not only on preferences for the general population, but also what is feasible and publicly acceptable for ongoing service delivery. For example, the most comprehensive panel size (“All conditions, regardless of severity”) introduces both ethical and operational issues. Ethically, it may be questionable whether adult-onset diseases should be the target of expanded carrier screening programs that seek to inform parents before birth.^20^ Operationally, this also introduces significant complexity: as more couples will test positive, their care decisions will likely require counselling and potentially treatment.^21^ Although respondents in our sample tended to prefer all conditions being included in the panel, the ramifications of providing such a comprehensive screening service may not have understood by participants. We strongly recommend that subsequent qualitative research focuses on the degree to which couples understand their reproductive options following receipt of a positive test, including the influence of included disorders on these decisions. Once couples are informed that they have tested positive for a condition, their options may include in-vitro fertilisation, prenatal testing, and adoption; if national screening is provided, the health system must also have available capacity for dealing with the consequences of positive tests.

In addition to potential respondent perceptions and understanding of screening, the purpose of carrier screening should also form a basis for how such programs will be adapted to future contexts. As gene therapies become more affordable, health service providers must decide what constitutes an acceptable level of intervention in preconception and prenatal care. As noted by Dive & Newson (2021), it may be beneficial to first anchor screening to principles of preventive care and reproductive autonomy, considering the arguments for which diseases justify inclusion in an iterative and holistic way based on factors beyond just prevention of affected births.^22^ These anchors will be useful to ensure that the principles of screening apply regardless of whether they are designed to detect a handful of severe diseases, or apply to the whole genome as it becomes increasingly better understood with regard to its impact on disease.

### Limitations

There are recurring limitations in expanded carrier screening programs throughout the literature that also apply to our study. The principle of disease severity is constantly evolving, especially as treatments become more effective and available,^23^ and prospective parents may have a different understanding of severity to a genetics specialist. The ethical quandaries regarding severity discussed above highlight that selecting genes for inclusion on a panel may be driven by practical concerns, including prevalence in the local population. However, they also introduce shifting operational concerns, including the probability of screening positive, which influences the need for genetic counselling and service capacity to deal with increased positive tests.

While we have attempted to put plausible distributions around cost parameters for the budget simulation in our model, a cost-effectiveness study would require a comprehensive micro-costing approach, including how different screening panels would be localised to the Singaporean context. The capability to provide screening with a 4-week or 8-week turnaround for no additional cost is a function of the laboratory capacity available to the national screening program; a sufficiently capable public or private sector can address these demands, but prices will likely shift as the prices of laboratory components and equipment, for example, also shift. Likewise, we budgeted for 25,000 eligible couples based on Singaporean childbirth rates; making screening available to premarital couples would substantially affect the budget impact, though it may better align the results of our experiment with revealed population preferences.

Discrete choice experiments are typically conducted to evaluate individual stated preferences, but reproductive choices are typically dyadic. Systematic reviews have reported on the preferences of couples for screening as a singular unit,^24^ however the decision-making process is complex, iterative, and typically difficult to map to a simple decision paradigm.^25^ We have addressed this by seeking out the theoretical boundaries of the dyadic uptake probability on an individual (partner 1 agrees OR partner 2 agrees) and shared (partner 1 agrees AND partner 2 agrees) basis, but as noted by Hershberger et al (2012) the reality is likely somewhere in between. A potential future study on how well stated individual preferences map to both stated dyadic preferences and revealed preferences can help expand upon this dynamic.

While we were interested in sociocultural factors that might affect preferences and uptake, including race/ethnicity, religion, and desire for procreation, our primary objective was to understand average preferences at the population level after accounting for individual preference heterogeneity. This was the purpose of the random effects specification. However, this may absorb potential heterogeneity at the subpopulation level, including for Malay and Indian participants. Our study was not designed to explore why preferences among racial or religious subgroups might differ, but we suggest that qualitative studies would be more suitable to understand whether expanded carrier screening programs may require further contextualisation based on language or shared cultural beliefs, especially as our study was conducted in English.

## CONCLUSIONS

We conducted a discrete choice experiment of population-level preferences for expanded carrier screening. Our results suggest that cost, number of genes included in the panel and wait times were the most important factors for Singaporeans of reproductive age when choosing whether or not to participate in a screening program, and we present predicted uptake and budget impact as a function of potential copayment amounts for testing to estimate the demand for screening.

## Data Availability

All data produced are available online at https://github.com/robinblythe/DCE_pop_screen

