## Supplement for "Designing national programs for expanded carrier screening: Results from a discrete-choice experiment in Singapore"

**Appendix 1: Survey instrument**

**A discrete choice experiment on general population preferences for a screening program for rare autosomal recessive disease risk in pre-conception couples**

Survey title: Survey on expanded carrier screening in Singapore

Logos:

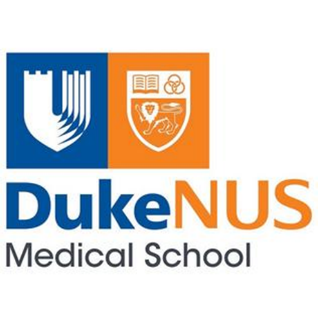

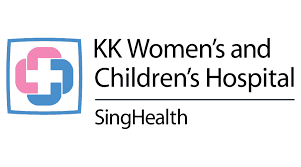

*Page 1: Introduction*

Thank you for taking part in this survey. We are a research team from Duke-NUS Medical School and KK Women’s and Children’s Hospital interested in understanding your preferences for carrier screening programs for rare inherited conditions. Your views will help us deliver a better service for all Singaporeans.

Carrier couple screening is intended for **all couples**, even if no one in their family is known to have a genetic condition. This is because carriers of genetic conditions and their relatives are typically healthy and do not have any symptoms of the conditions that they carry. So even if no one in your family is known to have a genetic condition, there is still a chance that you could be a carrier of a genetic condition.

A couple who is at **increased risk** for a particular condition has a 25% chance of having a pregnancy affected by the condition. This also means that the couple has a 75% chance of having a pregnancy that is unaffected by the condition.

*Page 2: Consent*

**Consent to participate**

I hereby acknowledge that by selecting “Yes”:

1. I agree to take part in this research study and that my responses can be used for research purposes.
2. I understand that I can choose to withdraw from this study at any time by choosing to close the browser window. However, my responses up to this point may still be used for research.
3. I will not benefit financially from this research.

RADIO BUTTONS: “Yes” OR “No”

IF RESPONSE TO CONSENT IS NO, PROCEED TO PAGE 4: END OF SURVEY

*Page 3*

What is your age?

INTEGER ENTRY, PROCEED TO END OF SURVEY IF AGE < 18 OR > 54

*Page 4*

1. With which of the following do you identify?
   a. Female
   b. Male
   c. Non-binary/third gender
   d. Prefer not to say

*Page 5*

2. What is your ancestry?
   a. Chinese
   b. Indian
   c. Malay
   d. Other

*Page 6*

3. What is your religious affiliation?
   a. Taoist
   b. Buddhist
   c. Hindu
   d. Muslim
   e. Catholic
   f. Christian
   g. No religious affiliation
   h. Other

*Page 7*

4. What is the highest level of education you have completed?
   a. PhD or Masters degree
   b. Bachelors degree
   c. Non-university higher education (e.g., polytechnic)
   d. Secondary school education
   e. Did not finish secondary school

*Page 8*

5. What is your marital status?
   a. Single
   b. In a relationship
   c. Married
   d. Divorced/widowed/separated

IF RESPONSE TO Q6 IS A OR D, PROCEED TO Q8

*Page 9*

6. Are you or your partner currently pregnant?
   a. Yes
   b. No

*Page 10*

7. Do you currently have children?
   a. Yes
   b. No

*Page 11*

8. Are you planning on having any children in the future?
   a. Yes
   b. No
   c. Unsure

*Page 12*

9. Do you, your children (if any), or someone you know have an inherited genetic condition, such as thalassaemia? Please select any that apply.
   a. Yes, I do
   b. Yes, my children do
   c. Yes, someone I know does
   d. No, not to my knowledge

*Page 13*

10. What is your level of agreement with the following statement:
    Genetic testing for inherited diseases should be available in Singapore.
    a. Strongly agree
    b. Agree
    c. Neutral
    d. Disagree
    e. Strongly disagree

IF RESPONSE TO Q11 IS D OR E, END SURVEY – GO TO PAGE 4

*End of demographic data collection*

*Page 14*

**INSTRUCTIONS ON HOW TO COMPLETE THIS SURVEY**

You will be shown two competing carrier screening programmes. Currently, some carrier screening programmes can cost up to $1,500 per couple for a comprehensive panel of conditions, not including additional costs for genetic counselling to understand the outcome of the test. Wait times can range up to several months to receive your results.

Imagine you are thinking of having children soon. You have heard about a new carrier screening service offered by the health service, and are thinking about whether you would like to participate in the programme. Please read through the options and decide which you prefer.

Below are the types of conditions detectable by expanded carrier screening in Singapore. Please note that each option includes the diseases above it. For example, Extremely severe and severe includes both disease groups that are considered extremely severe, as well as those that are considered severe. The option in green (All conditions regardless of severity) includes all diseases detectable by carrier screening.

1. **Extremely severe** (conditions with shortened lifespan in infancy/childhood or intellectual disability)
2. **Extremely severe and severe** (conditions with shortened lifespan in early adulthood, impaired mobility, or disabling organ impairment)
3. **Extremely severe, severe, and moderate** (conditions causing visual or hearing impairments and immune deficiency)
4. **All conditions regardless of severity**, including conditions with onset later in life such as genetic risks of heart disease or cancer

*DCE begins here. Format includes option 1, option 2, or an opt-out. Please see DCE design for the choice sets.*

END SURVEY

Thank you for participating in our survey! You may now close this window.

**Appendix 2: Localisation of systematic review and meta-analysis results to the Singaporean policy context and the think-aloud process**

*Willingness-to-pay and copayments*

Willingness-to-pay values in the meta-analysis were right-skewed, following a hurdle-Gamma distribution. When adapted to the Singaporean context, most of the posterior density corresponded to values that were insufficient to cover market prices for screening costs. To ensure that we covered a range that broadly represented incremental screening costs of different attribute levels, we chose the 10^th^, 50^th^, 80^th^, 95^th^, and 99^th^ percentiles of the posterior distribution and rounded them to easy-to-understand increments.

*Localisation to the Singaporean health system and existing healthcare infrastructure*

We adapted attribute levels from the review to the Singaporean context by engaging with genetics counsellors, clinical geneticists, health system administrators, and researchers to determine which aspects were relevant to the local health system. As we were also in the process of conducting a pilot study to understand patient preferences, the authors of the pilot (authors YB, MJY, JG, CC and SSJ) were able to identify preliminary beliefs from included patients regarding our attribute and level selections.

Based on the existing pilot program, we generally aimed to synchronise the results from the literature review with the pilot program, given much of the infrastructure was already developed. For example, genetics counsellors in Singapore were already trained to deliver carrier screening, so this was included as an option for clinician providing the screening service. Similarly, many Singaporeans access general practitioners and other primary care services through a multidisciplinary service (polyclinic), so we included a level for general practitioner/polyclinic. The relatively high certainty around capabilities of the pilot program helped to reduce the uncertainty in budget estimates for the extension of the pilot program.

*Validation: Think-aloud process*

We validated the survey tool (Appendix 1, above) by engaging three volunteers in think-aloud exercises. Volunteers were not paid, but agreed to review the survey tool as a personal favour to the lead author (RB). Two volunteers were research assistants at the lead author’s institution, and the third was a personal acquaintance and clinician. During the think-aloud process, volunteers were prompted to voice whatever thoughts came to them while completing the survey, and told that they could stop at any time to question or clarify. Volunteers agreed during the think-aloud exercise that all attributes and levels were simple to understand, with the exception of “Type of conditions screened” (*type*). We refined the attribute levels for *type* over several iterations to ensure that participants understood our intended meaning that each subsequent level of the attribute was a more comprehensive panel. We added text on mouseover to the survey to ensure that respondents could check the meaning of the level at any time. Our volunteers found this useful and did not believe there were any attributes or levels that were missing in the context of the Singaporean health system or local preferences.

*Comparison with census data*

As noted in the main text, we sought to ensure that our results were broadly representative of the Singapore population, according to 2025 census data. Census data were compiled using the SingStat Table Builder tool (<https://tablebuilder.singstat.gov.sg/>). Census comparisons were made prior to the pilot study design and then updated (where applicable) for Table 1 in the main text.

**Appendix 3: Experiment design**

*Choice experiment structure and sample size calculation*

The final choice experiment included seven attributes, which generated 4,860 (3^5^ x 5^1^ x 4^1^) possible profiles and 11,807,370 [4,860 × (4,860 – 1) / 2] possible pairwise combinations. Because a full factorial design was not feasible, a fractional factorial design was developed using Ngene software to generate a D-efficient design. The final design comprised 30 choice tasks, which were divided into three blocks of 10, so that each respondent completed 10 choice tasks. As no prior parameter estimates were available, small non-informative positive and negative priors were specified. Attribute levels were dummy coded.

Sample size requirements were informed by the *S_b_*-mean estimate, representing the minimum number of respondents required to achieve statistically significant parameter estimates at the 95% confidence level. The initial *S_b_*-mean estimates indicated a required sample size of approximately 15,000 respondents, which was not practically achievable. We sought a practical compromise between our budget constraints and the minimum established by Orme’s rule of thumb (500c/(tn)), where c is the maximum number of levels in any of the choice tasks, t is the number of tasks and n is the number of alternatives. As we had 5 levels in cost, 2 alternatives to choose from, and 10 tasks per respondent, this gave us a minimum of 125 respondents per block. Our budget allowed us to increase this to at least 160 respondents per block to ensure that we accounted for any potential loss of precision caused by respondents failing to make trade-offs, sometimes called the “garbage class.” We rounded this to an even 500 respondents.

*Survey delivery*

To limit the number of potential respondents who would always opt-out of screening, we pre-screened participants by including a question for whether participants agreed genetic carrier screening should be available in Singapore. We adopted a five-point Likert scale (Strongly agree, Agree, Neutral, Disagree, Strongly disagree) and excluded participants who answered Disagree or Strongly disagree. This accounted for approximately 5% of all respondents, who were screened out as per the design specified in Appendix 1.

We engaged a market research company, Kantar,^1^ to distribute our survey panel. Kantar applied internal checks for geolocation, completion speed, straight-lining (selecting the same option for all responses), and open-ended question validation.

**Appendix 4: Statistical model**

The model is described in Equations 1 through 3.

Equation 1 specifies the utility function, where utility $U_{njt}$ derived by individual *n* for alternative *j* in choice task *t* is a function of the vector of individual coefficients $\boldsymbol{\beta}_{\boldsymbol{n}}^{\top}$ and observed attributes $\boldsymbol{X}_{\boldsymbol{njt}}$. We used the *logitr* package for this analysis, which assumes the error term is Gumbel-distributed with mean 0 and variance $\sigma^{2}\left( \pi^{2}/6 \right)$.^2^

$$\begin{aligned} U_{njt}=\beta_{n}^{\top}X_{njt}+\varepsilon_{njt}\#\left( 1 \right) \end{aligned}$$

We specified random coefficients for preferences in Equation 2, assuming that preferences for each attribute were normally distributed and random parameters were independent. The ASC was also included as a random effect to capture potential heterogeneity in the choice to opt-out.

$$\begin{aligned} \beta_{n}\sim\mathcal{N}\left( \bar{\beta},\Sigma\right)\#\left( 2 \right) \end{aligned}$$

Equation 3 defines the mixed logit probability formula, where the following integral was approximated by simulation using 800 Sobol draws:

$$\begin{aligned} P_{nj}=\int\frac{\exp\left( \beta_{n}^{'}X_{njt} \right)}{\sum_{l} \exp\left( \beta_{n}^{'}X_{nlt} \right)}f\left( \beta_{n} \right)d\beta_{n}\#\left( 3 \right) \end{aligned}$$

Predicted uptake was calculated for each policy in comparison to opting-out, as if the policy was the only one on offer. It was then recalculated with all four policies in a single choice task, as if participants could choose a policy they found most preferable. Given that testing is a shared decision, we assessed two models of decision-making: individual (the couple would take the test if either one agreed) or shared (the couple would only take the test if both agreed). We calculated the shared decision as the square of the individual choice probability for individual policies vs the opt-out.

Finally, we also estimated uptake as a function of each level, varying one attribute at a time while holding all other attributes constant at their reference category. Uptake was calculated as a function of the five willingness-to-pay levels (Table S1).

*Table S1: Model coefficients from the final mixed multinomial logit.*

| Variable | Estimate | Std. Error | z-value | Pr(>\|z\|) |
| --- | --- | --- | --- | --- |
| cost_con | -0.003 | 0.000 | -13.141 | 0.000 |
| when_1 | 0.167 | 0.064 | 2.626 | 0.009 |
| when_2 | 0.075 | 0.075 | 0.994 | 0.320 |
| how_2 | 0.111 | 0.065 | 1.711 | 0.087 |
| how_3 | -0.050 | 0.070 | -0.710 | 0.478 |
| type_2 | 0.311 | 0.085 | 3.634 | 0.000 |
| type_3 | 0.497 | 0.082 | 6.023 | 0.000 |
| type_4 | 1.179 | 0.106 | 11.112 | 0.000 |
| edu_1 | 0.338 | 0.065 | 5.236 | 0.000 |
| edu_2 | 0.047 | 0.064 | 0.729 | 0.466 |
| clin_1 | -0.017 | 0.062 | -0.271 | 0.786 |
| clin_2 | 0.074 | 0.064 | 1.150 | 0.250 |
| wait_1 | 0.414 | 0.077 | 5.352 | 0.000 |
| wait_2 | 0.251 | 0.070 | 3.568 | 0.000 |
| asc | -5.109 | 0.390 | -13.085 | 0.000 |
| sd_cost_con | 0.003 | 0.000 | 15.478 | 0.000 |
| sd_when_1 | -0.440 | 0.108 | -4.082 | 0.000 |
| sd_when_2 | -0.549 | 0.114 | -4.801 | 0.000 |
| sd_how_2 | -0.268 | 0.156 | -1.719 | 0.086 |
| sd_how_3 | -0.242 | 0.166 | -1.451 | 0.147 |
| sd_type_2 | 0.303 | 0.158 | 1.915 | 0.055 |
| sd_type_3 | -0.270 | 0.177 | -1.526 | 0.127 |
| sd_type_4 | -0.978 | 0.117 | -8.376 | 0.000 |
| sd_edu_1 | 0.132 | 0.244 | 0.543 | 0.587 |
| sd_edu_2 | 0.199 | 0.176 | 1.128 | 0.259 |
| sd_clin_1 | 0.010 | 0.165 | 0.063 | 0.950 |
| sd_clin_2 | 0.238 | 0.146 | 1.623 | 0.105 |
| sd_wait_1 | -0.795 | 0.099 | -7.986 | 0.000 |
| sd_wait_2 | 0.104 | 0.241 | 0.430 | 0.667 |
| sd_asc | -4.567 | 0.345 | -13.232 | 0.000 |

*Table S2: Sensitivity analysis results when including interaction terms for Malay and Indian participants on cost and the alternative specific constant.*

| Variable | Estimate | Std. Error | z-value | Pr(>\|z\|) |
| --- | --- | --- | --- | --- |
| cost_con | -0.003 | 0.000 | -11.803 | 0.000 |
| cost_mal | 0.000 | 0.001 | -0.782 | 0.434 |
| cost_ind | 0.001 | 0.001 | 1.733 | 0.083 |
| when_1 | 0.182 | 0.065 | 2.810 | 0.005 |
| when_2 | 0.071 | 0.077 | 0.927 | 0.354 |
| how_2 | 0.104 | 0.066 | 1.573 | 0.116 |
| how_3 | -0.055 | 0.071 | -0.779 | 0.436 |
| type_2 | 0.315 | 0.086 | 3.680 | 0.000 |
| type_3 | 0.517 | 0.085 | 6.054 | 0.000 |
| type_4 | 1.195 | 0.111 | 10.764 | 0.000 |
| edu_1 | 0.343 | 0.066 | 5.180 | 0.000 |
| edu_2 | 0.055 | 0.066 | 0.841 | 0.400 |
| clin_1 | -0.013 | 0.063 | -0.209 | 0.835 |
| clin_2 | 0.064 | 0.065 | 0.983 | 0.325 |
| wait_1 | 0.437 | 0.079 | 5.523 | 0.000 |
| wait_2 | 0.252 | 0.072 | 3.491 | 0.000 |
| asc | -5.198 | 0.417 | -12.470 | 0.000 |
| asc_mal | -0.075 | 0.694 | -0.108 | 0.914 |
| asc_ind | -1.269 | 0.726 | -1.748 | 0.081 |
| sd_cost_con | 0.003 | 0.000 | 14.476 | 0.000 |
| sd_when_1 | -0.407 | 0.122 | -3.330 | 0.001 |
| sd_when_2 | -0.604 | 0.120 | -5.037 | 0.000 |
| sd_how_2 | 0.316 | 0.151 | 2.091 | 0.037 |
| sd_how_3 | 0.278 | 0.147 | 1.894 | 0.058 |
| sd_type_2 | 0.230 | 0.179 | 1.288 | 0.198 |
| sd_type_3 | 0.393 | 0.153 | 2.575 | 0.010 |
| sd_type_4 | -1.036 | 0.129 | -8.053 | 0.000 |
| sd_edu_1 | 0.269 | 0.149 | 1.804 | 0.071 |
| sd_edu_2 | 0.265 | 0.141 | 1.887 | 0.059 |
| sd_clin_1 | -0.026 | 0.126 | -0.206 | 0.837 |
| sd_clin_2 | -0.273 | 0.156 | -1.752 | 0.080 |
| sd_wait_1 | 0.819 | 0.102 | 8.062 | 0.000 |
| sd_wait_2 | 0.252 | 0.173 | 1.457 | 0.145 |
| sd_asc | -4.478 | 0.332 | -13.478 | 0.000 |

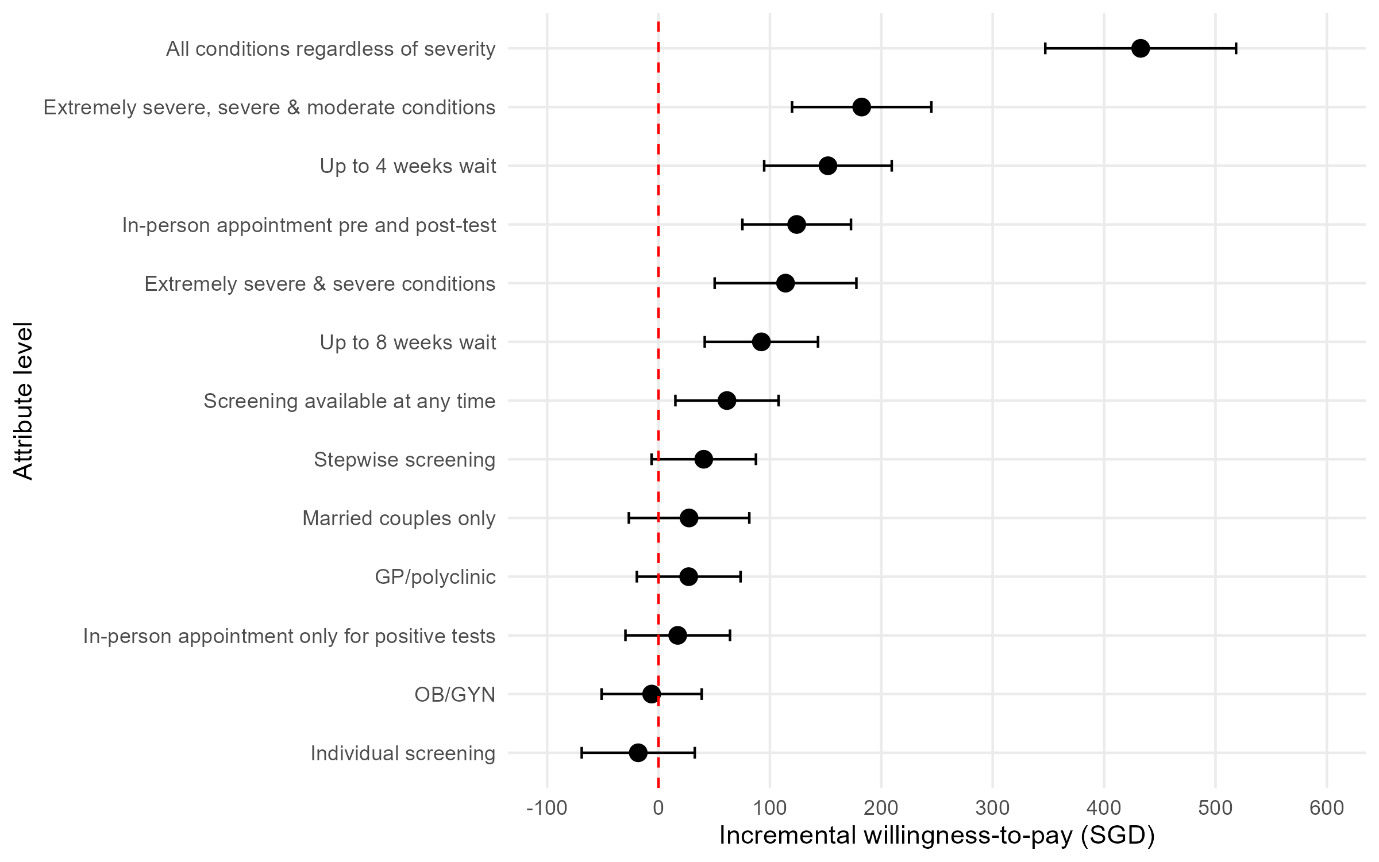

*Figure S1: Willingness-to-pay values for attribute levels relative to the reference group.*

*Budget impact and policy analysis*

Our three hypothetical policies, compared to the opt-out, were as follows:

- Policy 1: All attributes at the reference level. Screening is available for married couples pre-conception, with only couples’ results available and only extremely severe conditions included. Only online materials are available, and wait times are 16 weeks. The genetics counselling service handles screening.
- Policy 2: Extension of the existing pilot program. Screening is available for married couples pre-conception, with only couples’ results available, and both extremely severe and severe conditions are included. Online materials are available pre-testing but counselling is provided in the event of a positive test result and wait times are 8 weeks. The genetics counselling service handles screening.
- Policy 3: Based on estimated willingness-to-pay relative to the cost of service provision. Screening is available at any time, delivered in a stepwise manner (one partner screened first; if positive, second partner screened). Extremely severe, severe and moderate conditions are included. Counselling is available before and after testing and wait times are 4 weeks. The genetics counselling service handles screening.

We assumed that costs were roughly as follows at the policy level, based on operational data. As we were not able to obtain micro-costing, estimates were as follows: $430 per individual test for the pilot program (type level 2: extremely severe and severe), or $860 per couple. A smaller panel (type level 1: extremely severe only) was estimated to cost roughly half this amount, or $215 per individual/$430 per couple. To address uncertainty, this was sampled from a Normal distribution with mean $215 and standard deviation of $25. The cost of a single counselling visit was based on KK Women’s and Children’s Hospital (KKH) outpatient rates ($154.03), while the cost of an OB/GYN appointment was estimated at $240, based on internal KKH data.

At the time of analysis, type 3 (extremely severe, severe and moderate conditions) screening was available to Singaporeans by sending their samples to the US for approximately USD325 without accounting for shipping costs, converted to SGD at a rate of 1.28 SGD/USD, with exchange rates as of 18 March 2026 specified. To add uncertainty to the cost of locally conducting screening, we assumed that screening could be done locally based on a lognormal distribution with mean log($430) and standard deviation of log(1.11).

Stepwise screening was anticipated to cost less than couple screening based on the assumption that around 60% of individuals had at least one disease causing variant in the panel. The equation used was –(1 – 0.6) * C, where C was the cost of the test described above. This was sampled using a Beta distribution with shape parameters (60, 40). Finally, the rate of positive couples was estimated as a Beta distribution with parameters (1, 114). We direct readers to the code repository (<https://github.com/robinblythe/DCE_pop_screen>) for further detail.

**References used in Supplement**

1	Kantar. *Kantar Singapore*, <<https://www.kantar.com/locations/singapore#_>=> (2026).

2	Helveston, J. P. logitr: Fast estimation of multinomial and mixed logit models with preference space and willingness-to-pay space utility parameterizations. *Journal of Statistical Software* **105**, 1–37 (2023).
